# A scoping review protocol on *in vivo* human plastic exposure and health impacts

**DOI:** 10.1101/2022.02.10.22270706

**Authors:** Louise M. Goodes, Enoch V.S. Wong, Jennifer Alex, Louise Mofflin, Priyanka Toshniwal, Manuel Brunner, Terena Solomons, Emily White, Omrik Choudhury, Bhedita J. Seewoo, Yannick R. Mulders, Tristan Dale, Hamish J. Newman, Alina Naveed, Andrew B. Lowe, Delia V. Hendrie, Christos Symeonides, Sarah A. Dunlop

## Abstract

**Background:** Global plastic production has increased exponentially since the 1960s, with more than 6300 million metric tons of plastic waste generated to date. Studies have found a range of human health impacts associated with exposure to plastic chemicals. However, only a fraction of plastic chemicals used have been studied *in vivo*, and then often in animals, for acute toxicological effects. With many questions still unanswered about how long-term exposure to plastic impacts human health, there is an urgent need to map human *in vivo* research conducted to date, casting a broad net by searching terms for a comprehensive suite of plastic chemical exposures and the widest range of health domains.

**Methods:** This protocol describes a scoping review that will follow the recommended framework outlined in *2017 Guidance for the Conduct of Joanna Briggs Institute (JBI) Scoping Reviews*, to be reported in accordance with *Preferred Reporting Items for Systematic Reviews* and *Meta-Analyses extension for Scoping Reviews (PRISMA-ScR) Checklist*. A literature search of primary clinical studies in English from 1960 onwards will be conducted in MEDLINE (Ovid) and EMBASE (Ovid) databases. References eligible for inclusion will be identified through a quality-controlled, multi-level screening process. Extracted data will be presented in diagrammatic and tabular form, with a narrative summary addressing the review questions.

**Discussion:** This scoping review will comprehensively map the primary research undertaken to date on plastic exposure and human health. Secondary outputs will include extensive databases on plastic chemicals and human health outcomes/impacts.

**Registration:** Open Science Framework (OSF)-Standard Pre-Data Collection Registration, https://osf.io/gbxps DOI: 10.17605/OSF.IO/GBXPS

## Background

Continuous human exposure to plastic chemicals has become inevitable due to its ubiquity in the environment (1,2). ‘Plastic’ is a broad term for material made up of polymers and usually a number of chemical additives that impart desirable properties, such as flexibility and fire resistance (3,4). We will use the term ‘plastic materials’ as an umbrella term inclusive of plastic chemicals, plastic products and plastic particles; and ‘plastic chemicals’ to refer to polymers, monomers and/or additives.

As of 2017, approximately 8300 million metric tons (Mt) of virgin plastic chemicals had been produced, and as of 2015, 6300 million Mt of plastic waste generated (5). While some of the plastic waste is recycled or incinerated (5), the majority accumulates in landfills or in the environment (5), as macroplastic debris in the oceans (6–8), as microparticles in air (9–11), micro- or nanoparticles in soils (12–14) and in water supplies (15–18).

Plastic materials are used in every aspect of our lives (19) and common routes of exposure to plastic chemicals and particles (20) include inhalation (21), ingestion (including trophic transfer) (22–24), and direct dermal contact (25–27). Furthermore, children are exposed prenatally to plastic chemicals through maternal and paternal exposure (28), as well as postnatally through breast milk (29).

Of the thousands of plastic chemicals in use globally today, more than 20% have been identified as substances of concern by the European Union on the basis of persistence, bioaccumulation and/or toxicity, while 39% remain unclassified in terms of hazard (30). Some plastic chemicals are persistent organic pollutants that are known to bioaccumulate in human fatty tissues (31). A growing body of evidence links human exposure to various plastic chemicals with adverse health impacts (32–35) and public concern about the threat of plastic pollution on our health is high (36). Despite this, research efforts have focused on a few plastic chemicals for which established measurement techniques exist (37) and for which a limited range of health impacts have been identified – primarily endocrine disruption (38–40), reproductive toxicity (41,42), carcinogenicity (43,44), metabolic and nutritional conditions (45), and neurodevelopment (46,47). The findings of these studies are often inconsistent, as plastic chemical exposure can show sex-specific effects (38), differentially affect developmental stages (48), have health outcomes in association with specific plastic chemical mixtures (38), and can elicit non-monotonic responses (34). There is insufficient published data on the toxicity of micro- and nano-plastics in humans (49,50), however an absence of evidence of toxicity does not equate to evidence of safety (51).

We are not aware of any sources or publications comprehensively summarising research conducted to date on how plastic chemicals and plastic particles are impacting human health, or which populations, demographically and geographically, have been studied in this respect. Preliminary searches conducted in Prospero, Epistimonikos, Cochrane, Medline (Ovid) and Embase (Ovid) databases in May 2020 did not identify any previous or current scoping review on this broad topic. A number of systematic and literature reviews were identified, focusing on a small number of classes of plastic chemicals, such as phthalates, and bisphenol A (52–55), and certain diseases associated with these chemical species (56,57). These systematic and literature reviews addressed only a few specific chemicals and did not evaluate non disease-specific health domains, such as changes in gene expression, oxidative stress and the microbiome. Demographic and geographic bias (for example, due to social equity issues) were not evaluated, suggesting that many populations could be under-represented in the literature. For example, young children could be highly exposed and particularly vulnerable to the effects of some plastic chemicals (53,58), and populations in developing countries could be at increased exposure risk (59) due to a combination of higher plastic pollution burden (60), combined with inadequate waste management facilities and lower occupational health and safety regulations (61).

To demonstrate the scope of research conducted to date, and to inform future research efforts, we will conduct a scoping review to chart published primary human *in vivo* studies that investigated the relationship between exposure to plastic materials and human health since 1960. The scoping review aims to summarise the research to date, identify knowledge gaps and provide a framework for future studies in areas requiring more in-depth research.

### Review question

In primary human *in vivo* research conducted since 1960 on the effects of plastic materials on health, which plastic chemicals/particles and which health domains have (and have not) been investigated?

In relation to our main question, the following questions will inform sub-analyses:

1. Which populations (geographically, by age group, sex and general/special exposure risk status) have been examined?
2. Which study designs were adopted?
3. When were these studies conducted over time (Jan 1960–Jan 2021)?

### Inclusion and exclusion criteria

The Population, Exposure and Outcome (PEO) mnemonic (Table 1) was created to construct clear inclusion criteria

**Table 1:**
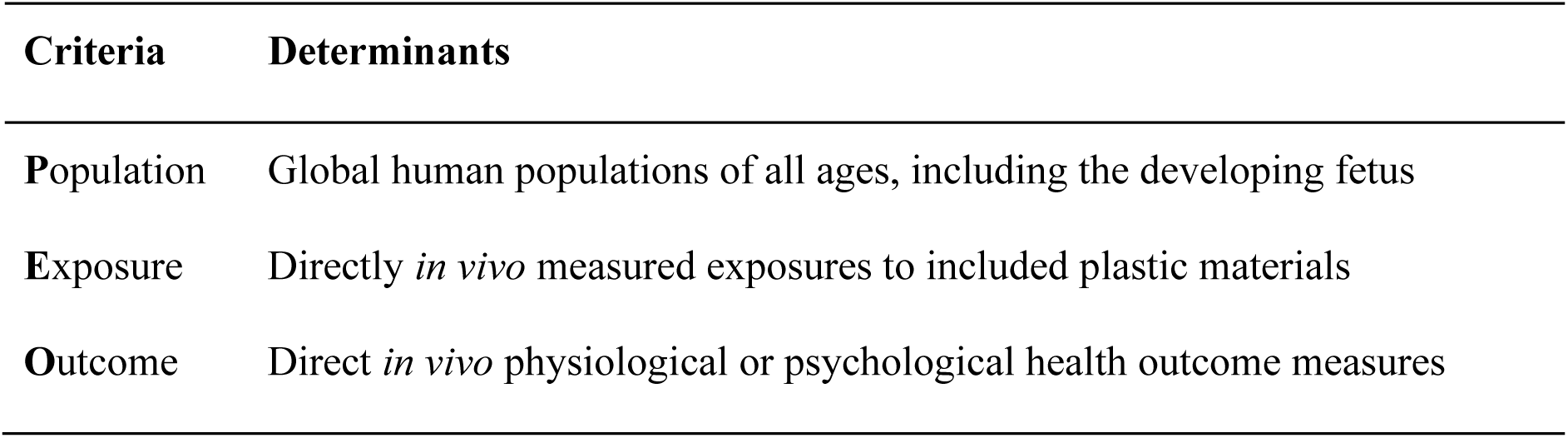
PEO framework for identifying the research question and inclusion criteria.

#### Population

The review will consider studies that investigate any population globally, and any age group, including the developing fetus. Animal studies and *in vitro* human studies will be excluded.

#### Exposure

Only *in vivo* directly measured exposures (i.e. via bio-samples, including for example blood, urine, breast milk and adipose tissue) of plastic materials that meet inclusion criteria will be included. All avenues of plastics exposure are considered relevant, including environmental contact via activities of everyday living, occupational exposure, accidental ingestion through contaminated food or water, cross-placental exposure, and ingestion through breast milk.

#### Plastics inclusion and exclusion criteria

This scoping review focuses on plastic materials to which humans are generally exposed, other than at the point of synthesis or product manufacturing. A systematic decision-making process (Figure 1) will be applied to determine the inclusion or exclusion of plastic chemicals. Briefly, plastic additives and synthetic polymers will be identified from preselected references: seven for additives (62–68), and seven for polymers (69–75). Chemicals that are classified as *additive – plasticiser, additive – flame retardant, per- and polyfluoroalkyl substances (PFAS), individual polymer* or *polymer class* and *bisphenols* will be included, unless commonly used in biomedical applications or as food additives (since inclusion would result in the retrieval of many thousands of references unrelated to the effects of plastic chemicals on health). Additional plastic chemicals identified during the screening process, or missing plastic chemicals that are structurally and systematically related to included substances (based on expert decision by Authors JA and MB, postdoctoral degrees in chemistry), will undergo the same selection criteria and subsequently included or excluded. Polymer metabolites, monomers other than bisphenols, and plastic additives other than plasticisers and flame retardants, are excluded from this review.

**Figure 1:** Schematic overview of the process for inclusion/exclusion of plastic chemicals in the scoping review

**Figure 2:** Study selection process

A comprehensive and searchable database of included plastic chemicals will be developed, indexing the multiple names, synonyms and commonly used terminology describing these chemicals.

#### Outcome

Only physiological and psychological changes in human health that are directly measured *in vivo* are included.

#### Human Health Outcome Measures

Health outcome measures include any assessment of change in structure or function of the human body, whether undertaken at the clinical, functional, physiological or cellular level. From preliminary searches, we anticipate encountering three types of health-related impact assessments in relation to plastics exposures:

1. Measures of direct impact on human health as a diagnosis of illness or disease, for example, breast cancer;
2. Clinical measures to detect a disease/disorder where no illness/disease has been diagnosed, for example oestrogen levels as a measure of change in ovarian function; and
3. Measures of impact on physiological or biochemical measures within an organ/body system that are not associated with a specific disorder, for example genetic and epigenetic changes and measures of metabolism and oxidative stress.

We will include measures of all the health-related impacts listed above, classifying them within the body system/organ in which they were investigated, using the hierarchical structure of the *International Classification of Disease 11*^*th*^ *Revision* (ICD-11) (76).

#### Types of sources

This scoping review considers peer-reviewed journal articles presenting primary human research, inclusive of all study designs. Excluded sources are literature/systematic reviews, meta-analyses, editorials, book reviews/chapters, perspectives, opinion/commentary pieces, conference abstracts and proceedings, and grey literature.

## Methods

### Design

The scoping review uses the methodological framework as described in the *Updated methodological guidance for the conduct of scoping reviews* (77) and reporting will be compliant with the *Preferred reporting items for systematic reviews and meta-analyses extension for scoping reviews (PRISMA-ScR) Checklist* (78) (see Additional file I for completed PRISMA-P checklist accompanying this Protocol) and informed by *Conducting high quality scoping reviews – challenges and solutions* (79).

### Constructing the search strategy

The search strategy aims to find published plastic chemicals/particles and human health studies from 1960 onwards. The starting point of 1960 was selected since, although large-scale use of plastic dates back to ∼1950, significant increases in global production occurred from the 1960s onward (5).

We identified key articles relevant for our scoping review by conducting an initial scoping search in Medline (Ovid) and Embase (Ovid) databases and examining the hierarchy of controlled vocabulary in these databases (MeSH and Emtree respectively). Indexed terms were selected representing plastic materials (including commonly used terms for chemical classes meeting our inclusion criteria) and a broad range of health outcome categories informed by preliminary searches and expert input from Author CS, a consultant paediatrician with relevant research expertise (most human health categories being represented, except for those obviously unrelated to plastic chemical exposure or likely to result in the retrieval of many irrelevant results). MeSH and Emtree terms, corresponding text words, text words contained in the titles and abstracts of key articles, and terms and synonyms for all specific plastic chemicals included in our database, will comprise the final search strategy, being developed in consultation with Research Librarian, Author TS. A large number of exclusion terms such as “plastic surgery”, plasticity, etc., will be utilised to minimise irrelevant results.

In summary, the search strategy will include: Health outcomes AND plastic material exposures NOT exclusions, limited to English language and dates ranging from 1960 to current. A search filter will be used in each database (Medline and Embase, via the Ovid platform) to identify human-only studies and exclude animal studies (80–82). The keywords and indexed terms representing health outcomes will be combined with Boolean operator OR and combined using the Boolean Operator AND with the keywords and indexed terms for plastic materials. The proximity operator ADJ will capture terms adjacent within a tested number of words within the title and abstract fields, and the truncation symbol * will be used where appropriate.

Scopus and Web of Science databases were investigated as sources, however due to the lack of controlled vocabulary and no filters for human-only studies, these will not be searched.

In anticipation of a very large number of results from this search, the reference lists of retrieved papers will not be reviewed. We will, however, review the reference lists associated with a related ‘Umbrella Review’ of systematic reviews with meta-analyses on this topic, being conducted by Minderoo Foundation and reported separately (*in preparation*).

### Study selection

All identified articles will be collated and uploaded into the DistillerSR (Evidence Partners, Ottawa, Canada) systematic review software application for screening and data extraction, and deduplicated. Relevant articles will then be selected via two screening stages, each preceded by an in-depth training process for the team of reviewers, who have postgraduate and postdoctoral degrees in chemistry, biochemistry, environmental and health sciences. Training will involve the provision of guidelines, including inclusion/exclusion criteria and practical examples of their application, education on classifying study design, access to our database of included plastic chemicals and demonstration of the screening process using relevant articles to be included for the scoping review, ensuring consistency and reliability of assessment decisions.

References will first be screened for relevance by title, abstract and index terms. Reviewers will initially screen several sets of the same 100 references and discuss their screening decisions, proceeding to single-screening after achieving an 85% level of agreement with subsequently agreed (resolved) group decisions over three sets. Articles with no abstract but potentially relevant to the scoping review will be included at the first screening stage. Auditing will be performed on a random selection of screened references to evaluate screening accuracy, and we will use the inbuilt artificial intelligence (DistillerAI) tool to identify possible false exclusions, re-screening references as necessary and thereby minimising the number of valuable articles being discarded.

During the second screening stage, full-text articles will be reviewed for eligibility. Training will again involve the screening of several sets of full-texts, with discussions after each set about screening decisions. Reviewers will proceed to single-screening only after achieving an 85% level of agreement with subsequently resolved group decisions and excluding no more than two eligible articles in error per 100 full-texts. Eligible articles will be included for data extraction, whereas articles excluded once will require a second reviewer to confirm the exclusion. Any disagreements will be resolved by a third reviewer with relevant experience.

### Data extraction

The data to be extracted will include specific publication details, study characteristics, relevant plastic exposures detected in the body and the health outcome measures analysed in relation to plastic exposures. Data extraction forms designed in DistillerSR (drafts are provided in Additional file II) will be piloted extensively during a period of training, involving independent reviewing followed by group meetings to discuss decisions and revise forms as necessary (final versions to be provided in the scoping review paper). Two reviewers will extract data from each full-text. Reviewer pairs will discuss conflicts that arise, with input from a third reviewer with the most relevant expertise if assistance is required to resolve decisions. Articles may be excluded during this phase if found ineligible or if it is impossible to extract data due to ambiguity or poor reporting quality, with the reasons for exclusion reported in the final paper.

The findings reported in the included studies, i.e. correlations between plastic exposures and health outcome measures, or lack thereof, will not be recorded, since we are following the typical scoping review framework, without conducting quality assessment of retrieved articles. The afore-mentioned ‘Umbrella Review’ of systematic reviews with meta-analyses will capture high-level evidence on this topic.

### Data analysis and presentation

Frequency counts and descriptive statistics will be presented for publication details and study characteristics. The extracted data will be presented graphically, in diagrammatic and tabular form, in a manner that aligns with the objective of this scoping review. A narrative summary will accompany the tabulated and charted results.

Graphical illustrations will show the trend, by year of publication, for when studies about the impact of plastics and health were conducted; the number of articles by country of authors and investigated study populations; age ranges and sex of participants; frequency of the different study designs used; the plastic chemical/particle exposures and health outcome measures. The results will be presented to address the research questions, make comparisons, identify methodologies used and identify research gaps.

## Discussion

This scoping review aims to provide a comprehensive overview of research published since 1960 on the exposure of humans to plastic materials and the health outcomes assessed. Due to the breadth of our review question, we anticipate a large number of studies to intersect with the PEO framework. A detailed search strategy has been designed to identify eligible studies based on all three criteria: *in vivo* human populations, directly measured plastic materials that are included according to clear criteria and scientifically reliable sources, and direct measures of human health outcomes.

While health outcomes traditionally involve diseases and health conditions, the potential impacts of plastic materials may be subtle, perhaps only becoming apparent over time. By adapting the ICD-11 sub-categories to include health outcomes not necessarily classified as disease states (e.g. epigenetic changes and oxidative stress measures), this scoping review aims to identify a wider suite of health-related outcome measures that should be taken into consideration for plastic exposure research. This database could also benefit future research endeavours.

In summary, this scoping review will provide an overview of the research so far undertaken on plastic material exposure and human health. This can serve as a basis to identify research clusters, by topic and geographical location of studied populations, as well as knowledge gaps, helping to prioritise specific areas in need of more in-depth research. Additionally, alongside the primary outputs, findings may be used to strengthen efforts to better regulate the production, importation and use of plastic materials that human populations are ubiquitously exposed to.

## Data Availability

All data produced in the present study are available upon reasonable request to the authors

## Additional (Supplementary) Information

Additional file I: Completed PRISMA-P checklist (PDF)

Additional file II: Draft data extraction forms (PDF)

## Abbreviations

ICD-11: *International Classification of Disease 11*^*th*^ *Revision*
JBI: *Joanna Briggs Institute*
OSF: *Open Science Framework*
PEO: *Population, Exposure and Outcome*
PRISMA-ScR: *Preferred reporting items for systematic reviews and meta-analyses extension for scoping reviews*

## DEFINITIONS

plastic chemicals: polymers, monomers and/or additives

plastic materials: plastic chemicals, plastic products and plastic particles

## DECLARATIONS

## Acknowledgements

The selection of specific plastic materials and associated terms was determined in consultation with polymer science expert Dr Marck Norret, The University of Western Australia. We also acknowledge the contribution from Brady Johnston, Minderoo Foundation, to create searchable online databases of our included plastics and health outcome measures, using the “Shiny” package in RStudio; and from Meredith Ryland and Joanne Webb, Minderoo Foundation, towards the writing of this manuscript.

## Authors’ contributions

*Scoping review design:* Sarah A. Dunlop, Christos Symeonides, Louise M. Goodes, Delia Hendrie, Tristan Dale, Hamish Newman, Enoch V.S. Wong, Priyanka Toshniwal *Manuscript:* Louise Goodes, Enoch V.S. Wong, Louise Mofflin, Priyanka Toshniwal, Manuel Brunner, Terena Solomons, Yannick Mulders, Sarah A. Dunlop (guarantor)

*Review of Manuscript:* Christos Symeonides, Jennifer Alex, Tristan Dale, Hamish Newman, Andrew Lowe, Delia Hendrie

*Identification of plastics and their sources:* Emily White, Jennifer Alex, Christos Symeonides, Andrew Lowe

*Classification of study designs, health outcomes and measures:* Bhedita Seewoo, Christos Symeonides, Louise Mofflin

*Coding and data management:* Omrik Choudhury, Enoch V.S. Wong

*Assistance with search strategy design:* Alina Naveed

## Availability of data and materials

All data generated or analysed will be included in the final published scoping review article and additional information related to the protocol will be available upon reasonable request.

## Ethics approval and consent to participate

Formal research ethics approval is not required for scoping review the published literature. The findings of the scoping review will be disseminated via Minderoo Foundation website, peer-reviewed scientific publication, presented at congresses or conferences, to policy makers and stakeholders aiming to achieve a world free of plastic pollution, and through various social media platforms. Data deposition and curation will be managed by Minderoo Foundation.

## Consent for publication

Not applicable.

