## Additional File II, Draft Data Extract Form for "A scoping review protocol on *in vivo* human plastic exposure and health impacts" (from another version of this preprint: version 1, 2022-02-10)

**DRAFT DATA EXTRACTION FORM**

1. Year of publication
2. Country of publication for first and last authors
3. Investigated population(s)
   a. Country/countries
   b. Individuals/pairs/families?
   c. Gender
   d. Age
   e. General/special risk (in terms of exposure)
4. Plastic exposures
5. Health outcome measures
6. Study design
7. Notes
